# Estimation of age and sex specific Glomerular Filtration Rate in the Abu Dhabi population and its association with mortality and Atherosclerotic cardiovascular outcome. A Retrospective Cohort Study

**DOI:** 10.1101/2024.08.10.24311788

**Authors:** Latifa Baynouna AlKetbi, Yousef Boobes, Nico Nagelkerke, Hamda Aleissaee, Noura AlShamsi, Mohammed AlMansoori, Ahmed Hemaid, Muna Jalal AlDobaee, Noura AlAlawi, Rudina Mubarak AlKetbi, Toqa Fahmawee, Basil AlHashaikeh, AlYazia AlAzeezi, Fatima Shuaib, Jawaher Alnuaimi, Esraa Mahmoud, Nayla AlAhbabi, Bachar Afandi

**Affiliations:** Ambulatory Healthcare Services; United Arab Emirates University; Seha Kidney Care - Tawam Hospital; Tawam Hospital, SEHA

**Keywords:** Risk Factors, Chronic Kidney Disease, Renal Hyperfiltration, secular trends, eGFR

## Abstract

The impact of abnormal Glomerular Filtration Rate (eGFR) on various adverse outcomes has been well studied; however, the United Arab Emirates (UAE), like many other regions in the world, remains understudied in this area.

**Method:** This retrospective cohort study estimates the age and sex-specific Glomerular Filtration Rate (eGFR) in the Abu Dhabi population and its association with mortality and Atherosclerotic cardiovascular (ASCVD) outcomes. The cohort of 8699 participants in a national cardiovascular disease screening from 2011 to 2013. The cohort was reevaluated in 2023 for mortality and cardiovascular outcomes. Reference eGFR percentiles were estimated from subjects without comorbidities using the LMS method.

**Results:** The reference percentiles of normal eGFR values showed a marked decrease with age, with small sex differences in the reference percentile distribution. A prognostic definition of renal hyperfiltration (RH) is suggested by the observation that subjects in the 97th percentile had a significantly higher incidence of ASCVD, although not statistically significant, in terms of mortality rate.

Older age, female sex, history of ASCVD, history of hypertension, being treated for hypertension, lower diastolic blood pressure, higher systolic blood pressure, lower HDL, higher HA1C, and higher vitamin D were significantly associated with lower eGFR percentiles. Subjects in the two categories within the RH range, the 95th and 97th percentiles, had a significantly higher prevalence of diabetes; they are older smokers with higher BMI, higher HA1C, higher HDL, lower vitamin D, and more likely to be males, with higher physical activity and have a lower prevalence of CHD.

**Conclusion:** The distribution of eGFR by age and sex is valuable for clinical decision-making in Abu Dhabi and likely for the Arab population in general. Although the 95th percentile of eGFR in this cohort showed a higher but nonsignificant risk, the 97th percentile is significantly associated with ASCVD, even more than subjects in the less than 10th eGFR percentile. This study provides important insights into the prevalence and risk factors associated with different eGFR percentiles in the Abu Dhabi population. The findings underscore the need for targeted interventions to address modifiable risk factors and prevent the progression of renal damage in this high-risk population.

## Introduction

Renal impairment adversely impacts patients’ health beyond kidney outcomes, such as end-stage renal disease (ESRD). It is strongly associated with mortality and atherosclerotic cardiovascular disease (ASCVD). (1, 2) (3). A meta-analysis of the association between non-dialysis-dependent CKD and the risk for all-cause and cardiovascular mortality demonstrated an exponential increase in absolute risk for death with decreasing kidney function even after adjustment for other established risk factors (4). There is also an increase in incidence and prevalence of cardiovascular events in patients with early CKD stages (CKD stages 1–3) compared with the general population, and patients with advanced CKD stages (CKD stages 4–5) exhibit a markedly elevated risk (4)Normal values for renal function and identifying abnormal cutoff points are critical due to their importance in management decisions. The CKD is classified by cause, the globular filtration rate (GFR) into five categories (G1–G5), and three albuminuria categories (A1–A3)(5). Reports have challenged the generalizability of current cutoff points and suggested multiple modifications to account for factors such as age, ethnicity, and other parameters (6) (7) (8), making the determination of “normal” ranges an active research area. Renal impairment is commonly identified with lower eGFR and the presence of proteinuria. Although higher GFR, or Renal Hyperfiltration (RH), is not yet classified as pathogenic in kidney disease classification, it has been associated with similar adverse outcomes (3, 9, 10). (11). There is also evidence that it precedes kidney damage and accelerates progression to CKD. Currently, RH is not classified as a disease state nor a risk factor recommended to be assessed and managed, and there are no established upper cutoff points between normal and diseased states (12). This is despite the evidence that it responds to newer medical interventions, such as renin-angiotensin-aldosterone system (RAAS) blockers and sodium-glucose cotransporter-2 inhibitors (SGLT2i) (13, 14). Therefore, targeting RH could influence the outcomes of many associated conditions.. such as diabetes, hypertension, obesity, and (15–19), as well as pregnancy and high protein intake (20, 21).

An important discussion is applying the Classification of CKD to different ethnicities, which requires more studies. The consensus now is that incorporating race in GFR estimation still faces challenges and that adjusting for ethnicity could lead to inaccurate estimation of eGFR due to genetic diversity within racial groups (8) (22).

No UAE or region studies have described the distribution of eGFR among the UAE or Arab population, and cutoff points based on adverse event occurrence are well needed. Misclassified as normal renal function, it will impact measures targeting risk factors. Additionally, many areas in the Arab world, especially the UAE, have high consanguinity,., which may constitute a unique population with unique characteristics.

This study aims to fill this gap by determining the UAE’s eGFR (ab)normal percentiles and associated risk factors and relating these percentiles to adverse outcomes. This research contributes to the global understanding of variation in estimated GFR levels. It underscores the importance of routine GFR assessments and the development of targeted interventions to prevent CKD and its complications.

## Method

This study included UAE nationals in the Emirate of Abu Dhabi. The Abu Dhabi Risk Study (ADRS) was a retrospective cohort study where subjects were randomly selected from participants in the cardiovascular prevention program, Weqaya (“prevention”), during 2011-2013 (23). This dataset was used to construct the eGFR percentiles for the healthy cohort population, as well as to study longitudinally outcomes reported during follow up years in relation to baseline, 2011-2013 characteristics. At two time points, different samples from the same national screening program were used. Participants of the screening in 2016-17 and in 2023-24 cohorts were studied to evaluate the change in the prevalence of eGFR percentiles distribution by age and sex.

Subjects in the first 2011-2013 cohort were reassessed in 2023, with a mean follow-up of 9.4 years. The ADRS included 8,699 subjects. Recorded variables included most conventional risk factors such as age, sex, Body Mass Index (BMI), HBA1C, Systolic Blood Pressure (SBP), Diastolic Blood Pressure (DBP), lipid profile (including total cholesterol and HDL), history of Chronic Kidney Diseases (CKD), diabetes, hypertension, and smoking. Additional factors included Estimated Glomerular Filtration Rate (eGFR) and Vitamin D levels. Follow-up data were collected by nurses and doctors as part of routine care. Baseline, Weqaya, assessments took place during 2011-2013, and in 2023, chart reviews were conducted by doctors and nurses to assess various outcomes of interest, such as cardiovascular events, diabetes, and other conditions. Detailed descriptions of the ADRS are available in earlier publications (24).

The method to determine the age-specific percentiles for normal eGFR was the LMS method (25) applied to individuals without known comorbidities, healthy. Excluded subjects from the eGFR percentile construction were those with diabetes, hypertension, cancer, heart disease, and stroke. The resultant age and sex-specific GFR percentiles are shown in Appendix 1.

The outcomes studied were death, coronary heart disease (CHD), and Atherosclerotic Cardiovascular diseases(ASCVD), either CHD or stroke. The relationship between eGFR percentiles and these outcomes was studied. While CKD diagnosis is clearly defined (5), hyperfiltration is not. Two definitions were studies in relation to the occurrence of adverse outcomes: an eGFR above the 95th percentiles (26) (27) and the National Kidney Foundation (NKF) criteria, with a GFR of 120 mL/min/1.73m^2^ or more regardless of age or sex (28).

To assess changes in prevalence, different participants in the Abu Dhabi preventive screening program in the years 2016-2017 and 2023-2024, a voluntary continuation of Weqaya screening, were assessed for changes in the prevalence of RH over the last decade. The 2016-2017 sample included 2554 subjects, and 2023-2024 included 10895 subjects. Both datasets were cross-sectional and included an electronic medical record extract report containing age, sex, BMI, and eGFR.

### Statistical analysis

The ADRS data was analyzed using the SPSS analysis program version 29. Descriptive statistics were used and presented as mean ± standard deviation (SD) for continuous variables and as percentages for categorical variables. Both binary logistic regression analysis and ordinal proportional odds models were used to explore the odds ratios (OR) of associations between RH and other baseline risk factors and their 95% C.I. The all-cause mortality risks and ASCVD events were estimated by multivariate Cox regression analysis adjusted for potentially confounding variables, including age, sex, body mass index (BMI), smoking status, HBA1C, history of DM, history of hypertension, vitamin D level, systolic and diastolic blood pressure, HDL, and total cholesterol. A significance level of p<0.05 was used throughout.

## Results

At baseline, subjects had a mean age of 38.8 years, BMI of 28.8, and eGFR of 111.3 mL/min/1.73m^2^. 16.3% of males reported current smoking, but smoking was rare among females). The diabetes prevalence was 22.2%, and the hypertension prevalence was 19.3% (Table 1).

In both sexes, a marked decrease with age of eGFR was observed, Table 2, Appendix 4. There also was a sex difference in the percentile distribution, with females losing GFR faster than males, clearly invalidating the idea of having a fixed age-sex independent cut-off value, notably 120 ml/min/1.73 m^2^, for hyperfiltration. The median of 121.9 ml/min/1.73 m^2^ for males and 126.97 ml/min/1.73 m^2^ among females at the age of 25 decreased to 96.38 ml/min/1.73 m^2^ and 95 ml/min/1.73 m^2^ at the age of 60 years in males and females, respectively. Classifying all cohort population based on the eGFR into the constructed percentiles showed that 33.8% of all males and 32.8% of all females are below the 50^th^ percentile.

The distribution of those over the 97^th^ percentile also differed between males and females. Among males, it was 4.2%, compared to 6.9% for females. Later, in the 2016-2017 cohort, the percentage in excess over expected percentiles increased. There were 17.5% of males exceeding the 97^th^ percentile compared to 7.4% of females. The 2023-2024 percentiles showed a similar distribution to 2016-2017, as shown in Table 2, with those having eGFR above the 97th percentile having a prevalence of 16.3% among males and 7% among females. If we consider hyperfiltration values of eGFR above the 95th percentile for age and sex, the 2011-2013 prevalence has nearly doubled among the total population, from 12.2% at baselines to 21.5% in 2016-2017 and 19.4% in 2023-2024 cohorts. Pregnancy was highly associated with RH (OR=8.6, 95% CI: 5.4-14, p < 0.001), and therefore, pregnant women were removed from the analysis as pregnancy is a transitional physiological status. Ordinal regression was used to investigate associations of extreme points of renal percentiles 3rd, 10th, 25th, and 95^th^ and 97^th^. As in Table 3-a, lower percentile eGFR percentiles studied with 3rd code =0, 10th-25th code =1, and 50th-75th are the reference. Table 3-b, higher percentiles eGFR, 50th -75th code =0, 95th code =1 and 97th as the reference. Significant associations existed between lower percentiles (a) and higher percentiles (b). All variables were data collected at baseline in 2011-2013. Older age, female sex, history of CHD, being on hypertension treatment, history of hypertension, higher levels of total cholesterol, and systolic blood pressure were associated with lower eGFR percentiles. There is a 4.4% increased risk of a low GFH for each additional year of age. History of hypertension at baseline increased this risk by 78%, and CHD risk at baseline increased by 57% among those with lower percentiles of eGFR. As well, there was a significant interaction between HBA1C level and diabetes with those with diabetes and worse control, higher HBA1C, having more risk of being in the lower eGFR percentile. Similarly, there was an interaction between age and diabetes with older diabetic patients significantly having higher risks. Surprisingly, Diabetes diagnosis was significantly associated with higher percentiles compared to lower, with non-diabetics having a 20.8% higher risk of being in the lower percentiles. Smoking was not significantly related to lower percentiles.

With regards to associations with higher percentiles, sex, and smoking status were not significantly related to higher percentiles. As well as younger age groups, lower levels of total cholesterol, lower levels of vitamin D, history of ASCVD, higher levels of HBA1C, higher levels of HDL, higher BMI, and less physically active individuals are at higher risk to be at higher percentiles categories. Table 3 For each one-unit increase in HBA1C, there is a 15.6% increase in the risk of being in a higher eGFR percentile. As well, for each one-unit drop of vitamin D, there is a 1.9% increased risk of being in the hyperfiltration range, i.e., 19% for every ten units; thus, a level of 20 has twice the risk of that of a person with a level of 70. HDL, on the other hand, showed a positive relationship with a 15.6% increase in risk for each unit, while lower total cholesterol levels were associated as well with a 22% higher unit decrease in levels.

Similarly, one unit increase in BMI was associated with a 1.2% increase in risk. There is a significant interaction between age and diabetes, with older diabetic patients being at higher risk of being in the hyperfiltration range. There is also more risk in that range with younger subjects with lower vitamin D levels.

During the decade of follow-up, death occurred among 39 females (1% of females) and 94 males (2.5% of males), and there were 62 ASCVD events among females (1.4% of females) and 239 events among males (5.5% of males). Coronary Artery Diseases occurred among 46 females (1.1% of females) and 204 males (4.7% of males). Using multivariate Cox regression, long-term effects of RH, which is based on the definition of eGFR of more than 120, showed only weak, non-significant associations with death, HR = 1.839 ( 0.827-4.088) P value=0.135 and coronary artery disease HR 2.058 (0.812-5.216) p-value 0.128. For ASCVD, HR=1.075 (0.465-2.487), p=0.87. However, in Cox survival analysis, there was a significant relationship between being above the 97^th^ percentile GR levels and the incidence of new CHD and new ASCVD, HR=1.65 (1.1-2.6), p=0.024 and HR=1.53 (1.005-2.4) p=0.048 respectively. This association with the 97th percentile was not significantly associated with mortality. Nevertheless, Kaplan-Meier analysis, as shown in Figure 3, shows a clear survival difference among the different percentile categories, with the 97^th^ percentile having the highest death rate among older males. The higher risk of the 97^th^ percentile is very clear and significant in the case of CHD and ASCVD.

## Discussion

This study reports the first eGFR percentiles for an Arab population derived from a community-based population health cohort. The eGFR levels are higher than those reported for Caucasian populations, such as data from Germany (29) (30), while they are closer to the normal values observed in Chinese populations (31). Similarly, in a 6-year longitudinal population-representative study from Delhi and Chennai, eGFR in India were higher than those reported in European cohorts (32). The 3rd percentile for an eGFR of less than 60, the CKD cutoff point, occurred at age 77 in males and 85 in females. These differences in eGFR between Asian and Caucasian populations warrant further study. Comorbidities and access to healthcare may be contributing factors. In the UAE, although healthcare access is among the best in the world, the prevalence of obesity, diabetes, prediabetes, and other CKD risk factors is very high (23). Furthermore, differences in body composition between Caucasians and other ethnicities, including Arabs, Indians, and Chinese, may also contribute to the variation. Lower muscle mass, which results in lower creatinine levels, could lead to an overestimation of true renal function (33)

The prevalence of renal hyperfiltration (RH) among the Abu Dhabi population is notably higher than that of other regions. In the United States, the highest reported RH prevalence among 12 to 29-year-old males was 11.8%, above the 95^th^ percentile, particularly among obese individuals (34). In Italy, among patients receiving specialist diabetes care in a large cohort, the prevalence was 3.4% (35), and in the general population of Korea, it was 2.69% (36). This study reported a higher prevalence of RH than these countries. RH is frequently observed in individuals with obesity, hypertension, and early-stage diabetes, serving as an early warning sign of potential kidney damage and progression to Chronic Kidney Disease (CKD) (37). The high prevalence observed in Abu Dhabi may be partly attributed to the high rates of obesity, smoking, diabetes, and other related risk factors, underscoring the need for interventions to address these contributing factors.

Renal Hyperfiltration (RH) is histologically characterized by glomerular hypertrophy, along with tubular hypertrophy and is associated with changes in intrarenal hemodynamic functions, including increased renal blood flow and intraglomerular hypertension. The increased intraglomerular pressure and glomerular hyperfiltration can lead to glomerulosclerosis and tubulointerstitial injury, eventually resulting in the deterioration of kidney function. Elevated glomerular pressure and hyperfiltration also contribute to endothelial dysfunction and vascular damage, which are key mechanisms in the development of CKD and Cardiovascular Disease (CVD) (28). These histological changes occur in response to various stimuli such as high-protein intake, hyperglycemia or insulin resistance, and obesity or metabolic syndrome (5, 12, 38). Additionally, RH is associated with an increase in albuminuria, which serves as an early marker of kidney damage and a predictor of cardiovascular events (39, 40).

Higher HBA1C levels and obesity were found to be associated with RH. This is supported by evidence from other studies, where even at prediabetes levels, higher HBA1C was associated with increased odds of having RH (41). In this study, diabetes diagnosis was not significantly associated with eGFR values lower than 10th percentiles compared to higher percentiles. Nevertheless, the interaction between diabetes and age and HBA1C levels indicates that the risk is higher in older and uncontrolled diabetics, whereas younger individuals with better control of diabetes, as represented by lower HBA1C, are associated with lower risk.

In patients with obesity, eGFR increases with body weight and is associated with the presence of albuminuria, indicating early kidney injury (42). These findings highlight the importance of early detection and intervention, including smoking cessation, in individuals with hyperfiltration to mitigate the risk of progressive kidney damage and improve long-term outcomes (43).

Although smoking status was not significantly associated with higher or lower eGFR percentiles in this study, the association between smoking and eGFR percentiles, despite the higher risk reported in other populations (13), warrants further investigation in this population. Maeda et al. found that smoking was linked to an elevated risk of glomerular hyperfiltration and dipstick proteinuria, emphasizing the renal impact of smoking (44, 45).

Improved management of modifiable risk factors, such as smoking cessation, better glycemic control, and weight management, may significantly reduce the negative health consequences of RH (35). Additionally, amelioration of hyperfiltration using ACE inhibitors (ACI) has been reported to significantly delay the long-term decline in GFR (46).

The association between hyperfiltration and lower vitamin D levels further suggests the need for early intervention strategies. Vitamin D deficiency has been implicated in the pathogenesis of CKD (47). Thus, vitamin D supplementation may hold promise in slowing CKD progression and preserving renal function (48). Moreover, emerging evidence suggests that vitamin D supplementation may exert renoprotective effects by attenuating inflammation, reducing proteinuria, and modulating the renin-angiotensin-aldosterone system (49, 50). Yet, further research is needed to elucidate the mechanisms underlying the association between hyperfiltration, vitamin D deficiency, and CKD progression as well as to evaluate the efficacy of vitamin D supplementation as an adjunctive therapy in CKD management (51). A counterintuitive finding that requires further investigation is the association of lower cholesterol, higher HDL, and lower SBP with hyperfiltration. Higher HDL-C levels were independently associated with accelerated GFR loss in a middle-aged nondiabetic population in a large cohort study in Norway (52). Whether high levels of HDL-C per se or higher levels of dysfunctional HDL-C contribute to endothelial dysfunction and vascular disease remains to be elucidated.

Since intrarenal RAAS activation plays a significant role in the pathogenesis of renal hyperfiltration, it is typically expected to be associated with higher blood pressure. (53)However, in our study, both a diagnosis of hypertension and higher blood pressure were associated with a lower risk of RH. The use of blood pressure medication was not a significant correlate of RH, but the potential role of RAAS inhibitors in this context warrants further research.

When comparing patients with RH to other subjects in terms of adverse outcomes, using eGFR 97th percentile as a predictor was significantly associated with CHD and ASCVD. Although a similar trend was observed in the Kaplan-Meier analysis, it was not statistically significant, likely due to the small number of deaths in this cohort. The significant association between CHD and ASCVD begins from the 90th percentile and becomes more pronounced at the 95th percentile, with the strongest association at the 97th percentile, particularly among males and older individuals. This prognostic cutoff point is clinically important for this population. In other Caucasian populations, the use of the 95th percentile might be attributed to previously described ethnic differences.

A higher incidence of ESRD amongst some ethnic minorities in the US has been attributed to a faster rate of GFR decline and more rapid progression of renal disease. In contrast, other ethnicities, such as Hispanics, exhibited lower mortality rates. These differences were hypothesized to be due to genetic variations in inflammatory or fibrotic pathways, renal response to injury, sensitivity to salt, toxins, and medications, hemodynamic autoregulation, or other factors involved in the progression of renal disease (54).

This study has several limitations that should be considered when interpreting its findings. The retrospective cohort design limits the ability to establish causality between identified risk factors and RH, as the associations observed do not necessarily imply direct cause-and-effect relationships. Additionally, the eGFR used to define RH may be influenced by inaccuracies in creatinine-based formulas, which can be affected by factors such as muscle mass, diet, and other individual characteristics. Unmeasured confounding variables, such as socioeconomic status, dietary habits, genetic predispositions, and other health behaviors, could also introduce potential bias. While the study included a large cohort, certain subgroups, particularly older age groups, had smaller sample sizes, which may limit the statistical power to detect significant associations. Despite these limitations, this study provides valuable information on eGFR values for this population and prognostic cutoff points based on the occurrence of adverse events. The findings underscore the need for further research on targeted interventions to address modifiable risk factors and prevent the progression of renal damage in this high-risk population.

## Data Availability

Data availability is restricted due to institution policies.

## Ethical approval and consent to participate

The study was approved by the Alain Human Ethics Committee, approval number 13/58, and Ambulatory Healthcare Services IRB 19-2022 and IRB 24. All methods were carried out under relevant guidelines and regulations. The authors confirm that the study was conducted in accordance with the Helsinki Declaration.

## Consent statement in the Ethics approval and consent to participate

Informed consent was waived by the IRBs as the study was designed for retrospective data gathered as part of patient care and anonymized at analysis.

## Authors’ contributions

LBK conceptualized the study, and LBK and NN analyzed data. LBK wrote the manuscript, and NN, BA, and YB reviewed the manuscript. All other co-authors collected data and reviewed the manuscript. All authors have read and approved the final manuscript.

## Acknowledgments

None.

## Consent to publish

Not Applicable.

## Availability of data and materials

Data availability is restricted due to institution policies.

---

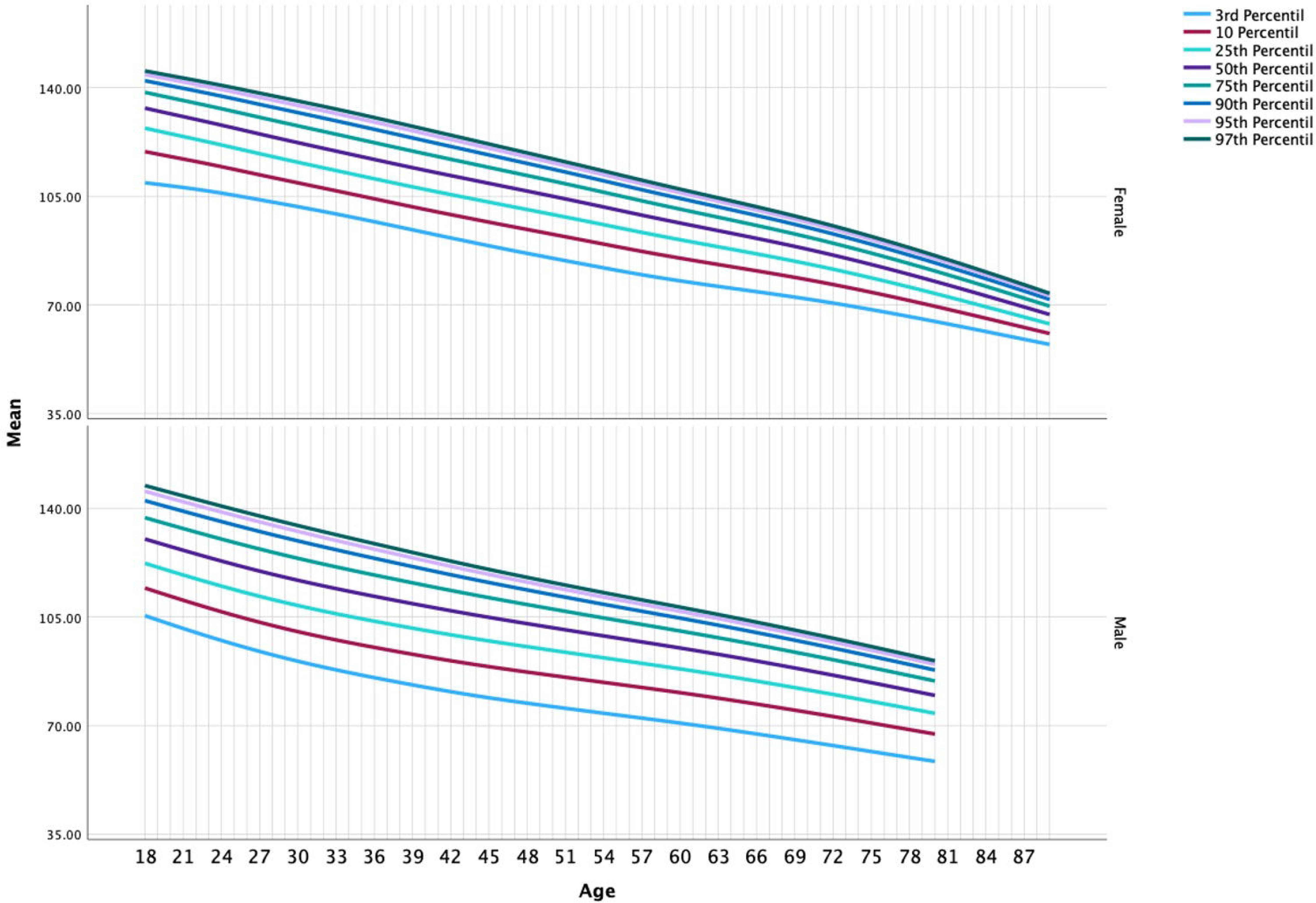

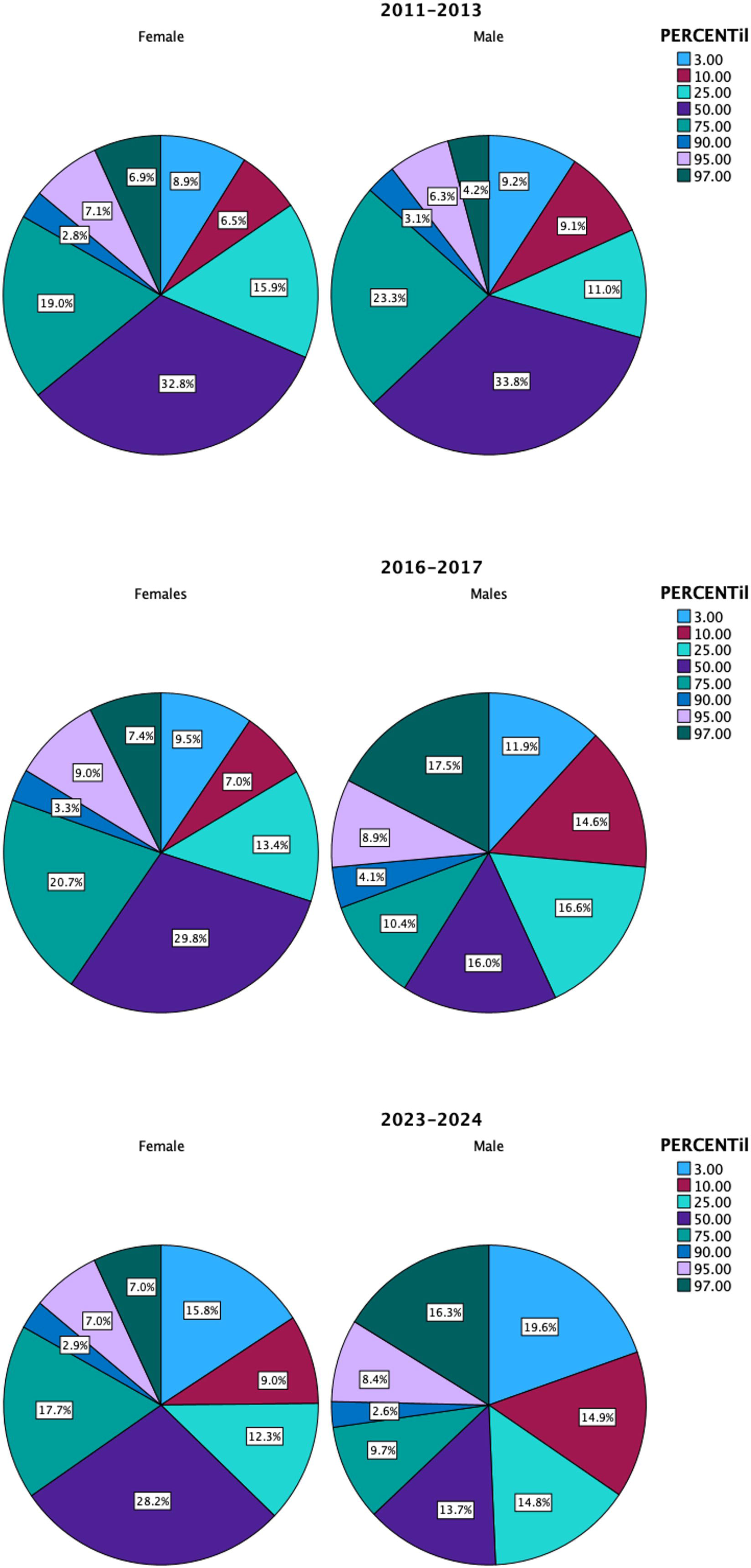

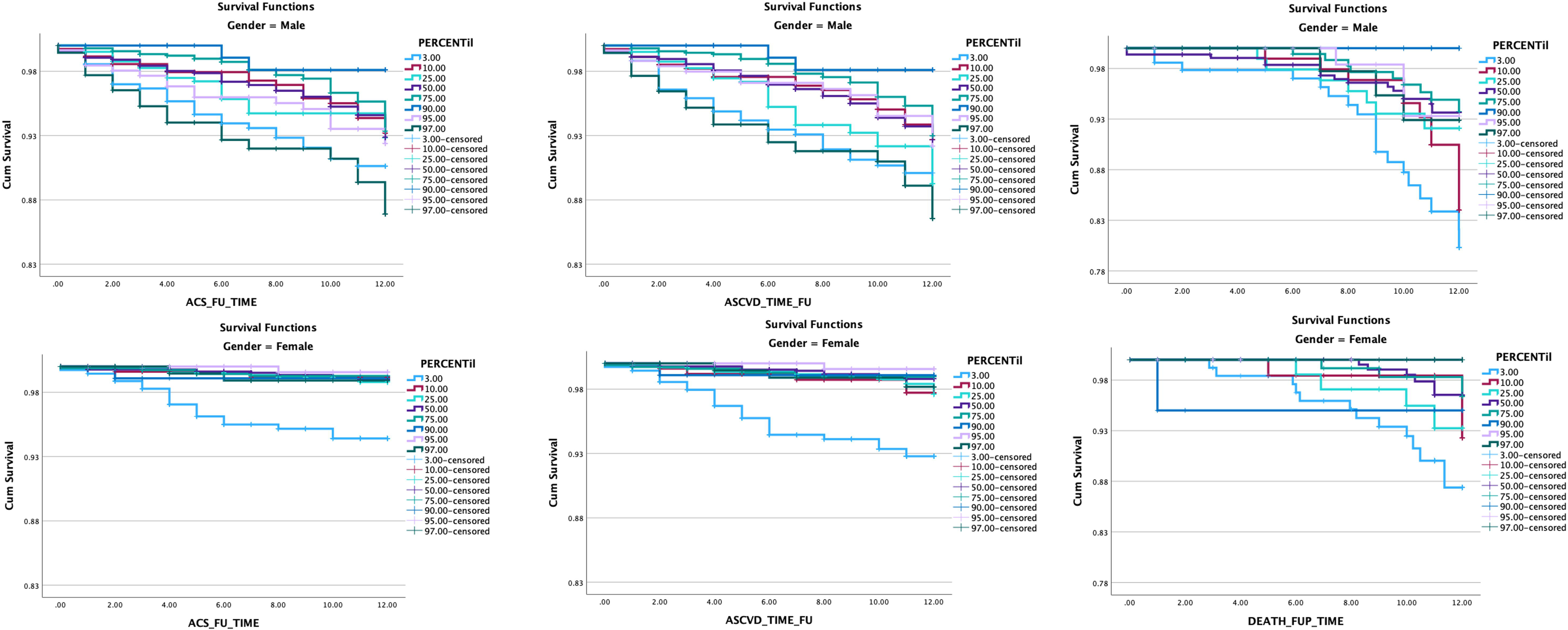

