## Supplementary material for "Estimation of age and sex specific Glomerular Filtration Rate in the Abu Dhabi population and its association with mortality and Atherosclerotic cardiovascular outcome. A Retrospective Cohort Study": table 3

Table 3 Associations of eGFR with studied risk factors

(a) lower percentile, eGFR percentiles, 3^rd^ code =0, 10^th^-25^th^ code =1, and 50^th^-75^th^ as the reference
(b) higher percentiles. eGFR percentiles, 50^th^ -75^th^ code =0, 95^th^ code =1 and 97^th^ as the reference
a. Ordinal regression of three levels of eGFR percentiles, 3^rd^, 10^th^-25^th^ and 50^th^-75^th^. The 97^th^ excluded.

|  |  | Estimate | P value | 95% CI | |
| --- | --- | --- | --- | --- | --- |
| Threshold | 3rd Percentile = 0.00 | -2.33 | <.001 | -3.407 | -1.253 |
|  | 10th-25th Percentile = 1.00] | -0.748 | 0.173 | -1.823 | 0.328 |
|  | Reference 50th, 75th and 90th Percentiles |  |  |  |  |
| Location | Cholesterol value | -0.096 | <.001 | -0.146 | -0.047 |
|  | HDL | 0.446 | <.001 | 0.285 | 0.606 |
|  | Vitamin D | -0.004 | <.001 | -0.007 | -0.002 |
|  | Intrction of age and diabetes | -0.013 | 0.014 | -0.024 | -0.003 |
|  | Age | 0.044 | <.001 | 0.025 | 0.063 |
|  | Square age | 0 | <.001 | -0.001 | 0 |
|  | SBP | -0.006 | 0.009 | -0.011 | -0.002 |
|  | DBP | 0.005 | 0.084 | -1.00E-03 | 0.012 |
|  | Intrction of HBA1C and diabetes | -0.067 | 0.051 | -0.134 | 0 |
|  | DM before screening=.00 | -1.208 | 0.002 | -1.956 | -0.46 |
|  | DM before screening=1.00 | 0a | . | . | . |
|  | ASCVD before=.00 | 0.429 | <.001 | 0.2 | 0.658 |
|  | ASCVD before=1.00 | 0a | . | . | . |
|  | Current Smoker=.00 | -0.108 | 0.252 | -0.292 | 0.077 |
|  | Current Smoker=1.00 | 0a | . | . | . |
|  | Sex=1.00] | -0.378 | <.001 | -0.488 | -0.268 |
|  | Sex=2.00] | 0a | . | . | . |
|  | On BP treatment=0] | 0.34 | 0.004 | 0.107 | 0.572 |
|  | On BP treatment=1] | 0a | . | . | . |
|  | [HTN before screening=.00] | 0.22 | 0.044 | 0.006 | 0.434 |
|  | [HTN before screening=1.00] | 0a | . | . | . |

b. Ordinal regression of three levels of eGFR percentiles, 97^th^, 95^th^, and 50^th^ -75^th^. the 3^rd^ 10^th^ and 25^th^ excluded.

|  |  | Estimate | P value | 95% CI | |
| --- | --- | --- | --- | --- | --- |
| Threshold | 50th and 75th Percentiles =0 .00 | 0.995 | 0.115 | -0.242 | 2.233 |
|  | 95th Percentile = 1.00 | 2.322 | <.001 | 1.082 | 3.562 |
|  | Reference is 97th Percentile |  |  |  |  |
| Location | Cholesterol value | -0.222 | <.001 | -0.295 | -0.149 |
|  | HBA1C | 0.156 | <.001 | 0.081 | 0.231 |
|  | HDL | 0.643 | <.001 | 0.447 | 0.838 |
|  | Vitamin D | -0.019 | <.001 | -0.03 | -0.008 |
|  | BMI | 0.012 | 0.03 | 0.001 | 0.022 |
|  | Intrction of age and diabetes | -0.016 | 0.018 | -0.029 | -0.003 |
|  | Age | -0.026 | 0.041 | -0.051 | -0.001 |
|  | Square age | 0 | 0.135 | -7.11E-05 | 0.001 |
|  | Intrction of age and vitamin D | 0 | 0.006 | 9.18E-05 | 0.001 |
|  | [Not active in physical activity =0] | 0.207 | 0.003 | 0.072 | 0.342 |
|  | [Active in physical activity=1] | 0a | . | . | . |
|  | DM before screening=.00 | -0.945 | 0.007 | -1.628 | -0.263 |
|  | DM before screening=1.00 | 0a | . | . | . |
|  | ASCVD before=.00 | 0.411 | 0.045 | 0.009 | 0.813 |
|  | ASCVD before=1.00 | 0a | . | . | . |
|  | Current Smoker=.00 | -0.179 | 0.122 | -0.406 | 0.048 |
|  | Current Smoker=1.00 | 0a | . | . | . |
