## Supplementary material for "Estimation of age and sex specific Glomerular Filtration Rate in the Abu Dhabi population and its association with mortality and Atherosclerotic cardiovascular outcome. A Retrospective Cohort Study": table 2

Table 2 Distribution of the eGFR percentiles by sex and age over among a three cohorts of screened population in Abu Dhabi

| **2011-2013** | | | | | | | | | | |
| --- | --- | --- | --- | --- | --- | --- | --- | --- | --- | --- |
|  |  | **P3** | **P10** | **P25** | **P50** | **P75** | **P90** | **P95** | **P97** | **total** |
| **Female** | <30 | 87.0 | 85.0 | 291.0 | 460.0 | 282.0 | 41.0 | 111.0 | 123.0 | 1480.0 |
|  |  | 0.1 | 0.1 | 0.2 | 0.3 | 0.2 | 0.0 | 0.1 | 0.1 | 1.0 |
|  | 30-39 | 78.0 | 66.0 | 186.0 | 385.0 | 246.0 | 33.0 | 73.0 | 85.0 | 1152.0 |
|  |  | 0.1 | 0.1 | 0.2 | 0.3 | 0.2 | 0.0 | 0.1 | 0.1 | 1.0 |
|  | 40-49 | 69.0 | 55.0 | 123.0 | 293.0 | 141.0 | 25.0 | 63.0 | 38.0 | 807.0 |
|  |  | 0.1 | 0.1 | 0.2 | 0.4 | 0.2 | 0.0 | 0.1 | 0.0 | 1.0 |
|  | 50-59 | 61.0 | 50.0 | 56.0 | 198.0 | 92.0 | 15.0 | 42.0 | 26.0 | 540.0 |
|  |  | 0.1 | 0.1 | 0.1 | 0.4 | 0.2 | 0.0 | 0.1 | 0.0 | 1.0 |
|  | 60-69 | 65.0 | 16.0 | 23.0 | 69.0 | 40.0 | 6.0 | 14.0 | 14.0 | 247.0 |
|  |  | 0.3 | 0.1 | 0.1 | 0.3 | 0.2 | 0.0 | 0.1 | 0.1 | 1.0 |
|  | 70-79 | 24.0 | 9.0 | 7.0 | 14.0 | 18.0 | 3.0 | 5.0 | 5.0 | 85.0 |
|  |  | 0.3 | 0.1 | 0.1 | 0.2 | 0.2 | 0.0 | 0.1 | 0.1 | 1.0 |
|  | >=80 | 3.0 | 2.0 | 2.0 | 2.0 | 1.0 | 0.0 | 0.0 | 6.0 | 16.0 |
|  |  | 0.2 | 0.1 | 0.1 | 0.1 | 0.1 | 0.0 | 0.0 | 0.4 | 1.0 |
|  | Total | 387.0 | 283.0 | 688.0 | 1421.0 | 820.0 | 123.0 | 308.0 | 297.0 | 4327.0 |
|  |  | 0.1 | 0.1 | 0.2 | 0.3 | 0.2 | 0.0 | 0.1 | 0.1 | 1.0 |
| **Male** | <30 | 90.0 | 96.0 | 162.0 | 495.0 | 300.0 | 46.0 | 81.0 | 52.0 | 1322.0 |
|  |  | 0.1 | 0.1 | 0.1 | 0.4 | 0.2 | 0.0 | 0.1 | 0.0 | 1.0 |
|  | 30-39 | 77.0 | 85.0 | 103.0 | 350.0 | 277.0 | 31.0 | 66.0 | 38.0 | 1027.0 |
|  |  | 0.1 | 0.1 | 0.1 | 0.3 | 0.3 | 0.0 | 0.1 | 0.0 | 1.0 |
|  | 40-49 | 48.0 | 76.0 | 73.0 | 223.0 | 190.0 | 23.0 | 44.0 | 34.0 | 711.0 |
|  |  | 0.1 | 0.1 | 0.1 | 0.3 | 0.3 | 0.0 | 0.1 | 0.0 | 1.0 |
|  | 50-59 | 52.0 | 59.0 | 72.0 | 222.0 | 133.0 | 22.0 | 45.0 | 24.0 | 629.0 |
|  |  | 0.1 | 0.1 | 0.1 | 0.4 | 0.2 | 0.0 | 0.1 | 0.0 | 1.0 |
|  | 60-69 | 87.0 | 53.0 | 46.0 | 137.0 | 83.0 | 8.0 | 23.0 | 26.0 | 463.0 |
|  |  | 0.2 | 0.1 | 0.1 | 0.3 | 0.2 | 0.0 | 0.1 | 0.1 | 1.0 |
|  | 70-79 | 42.0 | 23.0 | 22.0 | 38.0 | 28.0 | 4.0 | 16.0 | 7.0 | 180.0 |
|  |  | 0.2 | 0.1 | 0.1 | 0.2 | 0.2 | 0.0 | 0.1 | 0.0 | 1.0 |
|  | >=80 | 5.0 | 2.0 | 0.0 | 2.0 | 3.0 | 0.0 | 0.0 | 0.0 | 12.0 |
|  |  | 0.4 | 0.2 | 0.0 | 0.2 | 0.3 | 0.0 | 0.0 | 0.0 | 1.0 |
|  | Total | 401.0 | 394.0 | 478.0 | 1467.0 | 1014.0 | 134.0 | 275.0 | 181.0 | 4344.0 |
|  |  | 0.1 | 0.1 | 0.1 | 0.3 | 0.2 | 0.0 | 0.1 | 0.0 | 1.0 |
| **Total** | <30 | 177.0 | 181.0 | 453.0 | 955.0 | 582.0 | 87.0 | 192.0 | 175.0 | 2802.0 |
|  |  | 0.1 | 0.1 | 0.2 | 0.3 | 0.2 | 0.0 | 0.1 | 0.1 | 1.0 |
|  | 30-39 | 155.0 | 151.0 | 289.0 | 735.0 | 523.0 | 64.0 | 139.0 | 123.0 | 2179.0 |
|  |  | 0.1 | 0.1 | 0.1 | 0.3 | 0.2 | 0.0 | 0.1 | 0.1 | 1.0 |
|  | 40-49 | 117.0 | 131.0 | 196.0 | 516.0 | 331.0 | 48.0 | 107.0 | 72.0 | 1518.0 |
|  |  | 0.1 | 0.1 | 0.1 | 0.3 | 0.2 | 0.0 | 0.1 | 0.0 | 1.0 |
|  | 50-59 | 113.0 | 109.0 | 128.0 | 420.0 | 225.0 | 37.0 | 87.0 | 50.0 | 1169.0 |
|  |  | 0.1 | 0.1 | 0.1 | 0.4 | 0.2 | 0.0 | 0.1 | 0.0 | 1.0 |
|  | 60-69 | 152.0 | 69.0 | 69.0 | 206.0 | 123.0 | 14.0 | 37.0 | 40.0 | 710.0 |
|  |  | 0.2 | 0.1 | 0.1 | 0.3 | 0.2 | 0.0 | 0.1 | 0.1 | 1.0 |
|  | 70-79 | 66.0 | 32.0 | 29.0 | 52.0 | 46.0 | 7.0 | 21.0 | 12.0 | 265.0 |
|  |  | 0.2 | 0.1 | 0.1 | 0.2 | 0.2 | 0.0 | 0.1 | 0.0 | 1.0 |
|  | >=80 | 8.0 | 4.0 | 2.0 | 4.0 | 4.0 | 0.0 | 0.0 | 6.0 | 28.0 |
|  |  | 0.3 | 0.1 | 0.1 | 0.1 | 0.1 | 0.0 | 0.0 | 0.2 | 1.0 |
|  | Total | 788.0 | 677.0 | 1166.0 | 2888.0 | 1834.0 | 257.0 | 583.0 | 478.0 | 8671.0 |
|  |  | 0.1 | 0.1 | 0.1 | 0.3 | 0.2 | 0.0 | 0.1 | 0.1 | 1.0 |
| **2016-2017** | | | | | | | | | | |
|  |  | **P3** | **P10** | **P25** | **P50** | **P75** | **P90** | **P95** | **P97** |  |
| **Female** |  | 3.0 | 10.0 | 25.0 | 50.0 | 75.0 | 90.0 | 95.0 | 97.0 | total |
|  | <30 | 7.0 | 10.0 | 27.0 | 36.0 | 10.0 | 2.0 | 6.0 | 2.0 | 100.0 |
|  |  | 0.1 | 0.1 | 0.3 | 0.4 | 0.1 | 0.0 | 0.1 | 0.0 | 1.0 |
|  | 30-39 | 34.0 | 24.0 | 44.0 | 96.0 | 67.0 | 10.0 | 27.0 | 20.0 | 322.0 |
|  |  | 0.1 | 0.1 | 0.1 | 0.3 | 0.2 | 0.0 | 0.1 | 0.1 | 1.0 |
|  | 40-49 | 21.0 | 20.0 | 38.0 | 132.0 | 69.0 | 11.0 | 16.0 | 14.0 | 321.0 |
|  |  | 0.1 | 0.1 | 0.1 | 0.4 | 0.2 | 0.0 | 0.1 | 0.0 | 1.0 |
|  | 50-59 | 31.0 | 23.0 | 38.0 | 73.0 | 73.0 | 10.0 | 30.0 | 23.0 | 301.0 |
|  |  | 0.1 | 0.1 | 0.1 | 0.2 | 0.2 | 0.0 | 0.1 | 0.1 | 1.0 |
|  | 60-69 | 18.0 | 11.0 | 16.0 | 33.0 | 35.0 | 6.0 | 24.0 | 16.0 | 159.0 |
|  |  | 0.1 | 0.1 | 0.1 | 0.2 | 0.2 | 0.0 | 0.2 | 0.1 | 1.0 |
|  | 70-79 | 6.0 | 0.0 | 3.0 | 1.0 | 6.0 | 2.0 | 10.0 | 12.0 | 40.0 |
|  |  | 0.2 | 0.0 | 0.1 | 0.0 | 0.2 | 0.1 | 0.3 | 0.3 | 1.0 |
|  | >=80 | 3.0 | 0.0 | 2.0 | 3.0 | 0.0 | 0.0 | 0.0 | 6.0 | 14.0 |
|  |  | 0.2 | 0.0 | 0.1 | 0.2 | 0.0 | 0.0 | 0.0 | 0.4 | 1.0 |
|  | Total | 120.0 | 88.0 | 168.0 | 374.0 | 260.0 | 41.0 | 113.0 | 93.0 | 1257.0 |
| **Male** |  | 0.1 | 0.1 | 0.1 | 0.3 | 0.2 | 0.0 | 0.1 | 0.1 | 1.0 |
|  | <30 | 28.0 | 17.0 | 25.0 | 21.0 | 12.0 | 7.0 | 9.0 | 24.0 | 143.0 |
|  |  | 0.2 | 0.1 | 0.2 | 0.1 | 0.1 | 0.0 | 0.1 | 0.2 | 1.0 |
|  | 30-39 | 47.0 | 73.0 | 83.0 | 76.0 | 47.0 | 21.0 | 48.0 | 86.0 | 481.0 |
|  |  | 0.1 | 0.2 | 0.2 | 0.2 | 0.1 | 0.0 | 0.1 | 0.2 | 1.0 |
|  | 40-49 | 36.0 | 56.0 | 68.0 | 56.0 | 36.0 | 10.0 | 26.0 | 53.0 | 341.0 |
|  |  | 0.1 | 0.2 | 0.2 | 0.2 | 0.1 | 0.0 | 0.1 | 0.2 | 1.0 |
|  | 50-59 | 19.0 | 24.0 | 13.0 | 25.0 | 22.0 | 9.0 | 20.0 | 27.0 | 159.0 |
|  |  | 0.1 | 0.2 | 0.1 | 0.2 | 0.1 | 0.1 | 0.1 | 0.2 | 1.0 |
|  | 60-69 | 14.0 | 12.0 | 20.0 | 20.0 | 14.0 | 4.0 | 6.0 | 21.0 | 111.0 |
|  |  | 0.1 | 0.1 | 0.2 | 0.2 | 0.1 | 0.0 | 0.1 | 0.2 | 1.0 |
|  | 70-79 | 8.0 | 4.0 | 2.0 | 6.0 | 2.0 | 2.0 | 5.0 | 12.0 | 41.0 |
|  |  | 0.2 | 0.1 | 0.0 | 0.1 | 0.0 | 0.0 | 0.1 | 0.3 | 1.0 |
|  | >=80 | 0.0 | 0.0 | 1.0 | 0.0 | 0.0 | 0.0 | 0.0 | 1.0 | 2.0 |
|  |  | 0.0 | 0.0 | 0.5 | 0.0 | 0.0 | 0.0 | 0.0 | 0.5 | 1.0 |
|  | Total | 152.0 | 186.0 | 212.0 | 204.0 | 133.0 | 53.0 | 114.0 | 224.0 | 1278.0 |
| **Total** |  | 0.1 | 0.1 | 0.2 | 0.2 | 0.1 | 0.0 | 0.1 | 0.2 | 1.0 |
|  | <30 | 35.0 | 27.0 | 52.0 | 57.0 | 22.0 | 9.0 | 15.0 | 26.0 | 243.0 |
|  |  | 0.1 | 0.1 | 0.2 | 0.2 | 0.1 | 0.0 | 0.1 | 0.1 | 1.0 |
|  | 30-39 | 81.0 | 97.0 | 127.0 | 172.0 | 114.0 | 31.0 | 75.0 | 106.0 | 803.0 |
|  |  | 0.1 | 0.1 | 0.2 | 0.2 | 0.1 | 0.0 | 0.1 | 0.1 | 1.0 |
|  | 40-49 | 57.0 | 76.0 | 106.0 | 188.0 | 105.0 | 21.0 | 42.0 | 67.0 | 662.0 |
|  |  | 0.1 | 0.1 | 0.2 | 0.3 | 0.2 | 0.0 | 0.1 | 0.1 | 1.0 |
|  | 50-59 | 50.0 | 47.0 | 51.0 | 98.0 | 95.0 | 19.0 | 50.0 | 50.0 | 460.0 |
|  |  | 0.1 | 0.1 | 0.1 | 0.2 | 0.2 | 0.0 | 0.1 | 0.1 | 1.0 |
|  | 60-69 | 32.0 | 23.0 | 36.0 | 53.0 | 49.0 | 10.0 | 30.0 | 37.0 | 270.0 |
|  |  | 0.1 | 0.1 | 0.1 | 0.2 | 0.2 | 0.0 | 0.1 | 0.1 | 1.0 |
|  | 70-79 | 14.0 | 4.0 | 5.0 | 7.0 | 8.0 | 4.0 | 15.0 | 24.0 | 81.0 |
|  |  | 0.2 | 0.0 | 0.1 | 0.1 | 0.1 | 0.0 | 0.2 | 0.3 | 1.0 |
|  | >=80 | 3.0 | 0.0 | 3.0 | 3.0 | 0.0 | 0.0 | 0.0 | 7.0 | 16.0 |
|  |  | 0.2 | 0.0 | 0.2 | 0.2 | 0.0 | 0.0 | 0.0 | 0.4 | 1.0 |
|  | Total | 272.0 | 274.0 | 380.0 | 578.0 | 393.0 | 94.0 | 227.0 | 317.0 | 2535.0 |
|  |  | 0.1 | 0.1 | 0.2 | 0.2 | 0.2 | 0.0 | 0.1 | 0.1 | 1.0 |
| **2023-2024** | | | | | | | | | | |
|  | **2023-2024** | **P3** | **P10** | **P25** | **P50** | **P75** | **P90** | **P95** | **P97** | **total** |
| **Female** | <30 | 386.0 | 284.0 | 568.0 | 1093.0 | 585.0 | 91.0 | 203.0 | 181.0 | 3391.0 |
|  |  | 0.1 | 0.1 | 0.2 | 0.3 | 0.2 | 0.0 | 0.1 | 0.1 | 1.0 |
|  | 30-39 | 445.0 | 311.0 | 486.0 | 1119.0 | 728.0 | 142.0 | 322.0 | 331.0 | 3884.0 |
|  |  | 0.1 | 0.1 | 0.1 | 0.3 | 0.2 | 0.0 | 0.1 | 0.1 | 1.0 |
|  | 40-49 | 565.0 | 344.0 | 468.0 | 1157.0 | 651.0 | 93.0 | 234.0 | 183.0 | 3695.0 |
|  |  | 0.2 | 0.1 | 0.1 | 0.3 | 0.2 | 0.0 | 0.1 | 0.1 | 1.0 |
|  | 50-59 | 398.0 | 249.0 | 211.0 | 654.0 | 389.0 | 51.0 | 122.0 | 124.0 | 2198.0 |
|  |  | 0.2 | 0.1 | 0.1 | 0.3 | 0.2 | 0.0 | 0.1 | 0.1 | 1.0 |
|  | 60-69 | 349.0 | 145.0 | 112.0 | 306.0 | 280.0 | 44.0 | 138.0 | 96.0 | 1470.0 |
|  |  | 0.2 | 0.1 | 0.1 | 0.2 | 0.2 | 0.0 | 0.1 | 0.1 | 1.0 |
|  | 70-79 | 228.0 | 59.0 | 61.0 | 62.0 | 123.0 | 32.0 | 63.0 | 95.0 | 723.0 |
|  |  | 0.3 | 0.1 | 0.1 | 0.1 | 0.2 | 0.0 | 0.1 | 0.1 | 1.0 |
|  | >=80 | 100.0 | 16.0 | 19.0 | 17.0 | 9.0 | 0.0 | 15.0 | 77.0 | 253.0 |
|  |  | 0.4 | 0.1 | 0.1 | 0.1 | 0.0 | 0.0 | 0.1 | 0.3 | 1.0 |
|  | Total | 2471.0 | 1408.0 | 1925.0 | 4408.0 | 2765.0 | 453.0 | 1097.0 | 1087.0 | 15614.0 |
|  |  | 0.2 | 0.1 | 0.1 | 0.3 | 0.2 | 0.0 | 0.1 | 0.1 | 1.0 |
| **Male** | <30 | 599.0 | 465.0 | 438.0 | 491.0 | 385.0 | 116.0 | 327.0 | 552.0 | 3373.0 |
|  |  | 0.2 | 0.1 | 0.1 | 0.1 | 0.1 | 0.0 | 0.1 | 0.2 | 1.0 |
|  | 30-39 | 702.0 | 606.0 | 618.0 | 640.0 | 412.0 | 119.0 | 350.0 | 649.0 | 4096.0 |
|  |  | 0.2 | 0.1 | 0.2 | 0.2 | 0.1 | 0.0 | 0.1 | 0.2 | 1.0 |
|  | 40-49 | 637.0 | 580.0 | 639.0 | 514.0 | 383.0 | 79.0 | 330.0 | 619.0 | 3781.0 |
|  |  | 0.2 | 0.2 | 0.2 | 0.1 | 0.1 | 0.0 | 0.1 | 0.2 | 1.0 |
|  | 50-59 | 514.0 | 389.0 | 378.0 | 331.0 | 226.0 | 70.0 | 207.0 | 452.0 | 2567.0 |
|  |  | 0.2 | 0.2 | 0.1 | 0.1 | 0.1 | 0.0 | 0.1 | 0.2 | 1.0 |
|  | 60-69 | 358.0 | 249.0 | 241.0 | 186.0 | 120.0 | 30.0 | 104.0 | 236.0 | 1524.0 |
|  |  | 0.2 | 0.2 | 0.2 | 0.1 | 0.1 | 0.0 | 0.1 | 0.2 | 1.0 |
|  | 70-79 | 251.0 | 113.0 | 91.0 | 57.0 | 46.0 | 5.0 | 40.0 | 111.0 | 714.0 |
|  |  | 0.4 | 0.2 | 0.1 | 0.1 | 0.1 | 0.0 | 0.1 | 0.2 | 1.0 |
|  | >=80 | 144.0 | 24.0 | 17.0 | 22.0 | 16.0 | 4.0 | 10.0 | 38.0 | 275.0 |
|  |  | 0.5 | 0.1 | 0.1 | 0.1 | 0.1 | 0.0 | 0.0 | 0.1 | 1.0 |
|  | Total | 3205.0 | 2426.0 | 2422.0 | 2241.0 | 1588.0 | 423.0 | 1368.0 | 2657.0 | 16330.0 |
|  |  | 0.2 | 0.1 | 0.1 | 0.1 | 0.1 | 0.0 | 0.1 | 0.2 | 1.0 |
| **Total** | <30 | 985.0 | 749.0 | 1006.0 | 1584.0 | 970.0 | 207.0 | 530.0 | 733.0 | 6764.0 |
|  |  | 0.1 | 0.1 | 0.1 | 0.2 | 0.1 | 0.0 | 0.1 | 0.1 | 1.0 |
|  | 30-39 | 1147.0 | 917.0 | 1104.0 | 1759.0 | 1140.0 | 261.0 | 672.0 | 980.0 | 7980.0 |
|  |  | 0.1 | 0.1 | 0.1 | 0.2 | 0.1 | 0.0 | 0.1 | 0.1 | 1.0 |
|  | 40-49 | 1202.0 | 924.0 | 1107.0 | 1671.0 | 1034.0 | 172.0 | 564.0 | 802.0 | 7476.0 |
|  |  | 0.2 | 0.1 | 0.1 | 0.2 | 0.1 | 0.0 | 0.1 | 0.1 | 1.0 |
|  | 50-59 | 912.0 | 638.0 | 589.0 | 985.0 | 615.0 | 121.0 | 329.0 | 576.0 | 4765.0 |
|  |  | 0.2 | 0.1 | 0.1 | 0.2 | 0.1 | 0.0 | 0.1 | 0.1 | 1.0 |
|  | 60-69 | 707.0 | 394.0 | 353.0 | 492.0 | 400.0 | 74.0 | 242.0 | 332.0 | 2994.0 |
|  |  | 0.2 | 0.1 | 0.1 | 0.2 | 0.1 | 0.0 | 0.1 | 0.1 | 1.0 |
|  | 70-79 | 479.0 | 172.0 | 152.0 | 119.0 | 169.0 | 37.0 | 103.0 | 206.0 | 1437.0 |
|  |  | 0.3 | 0.1 | 0.1 | 0.1 | 0.1 | 0.0 | 0.1 | 0.1 | 1.0 |
|  | >=80 | 244.0 | 40.0 | 36.0 | 39.0 | 25.0 | 4.0 | 25.0 | 115.0 | 528.0 |
|  |  | 0.5 | 0.1 | 0.1 | 0.1 | 0.0 | 0.0 | 0.0 | 0.2 | 1.0 |
|  | Total | 5676.0 | 3834.0 | 4347.0 | 6649.0 | 4353.0 | 876.0 | 2465.0 | 3744.0 | 31944.0 |
|  |  | 0.2 | 0.1 | 0.1 | 0.2 | 0.1 | 0.0 | 0.1 | 0.1 | 1.0 |
