## Supplementary material for "Estimation of age and sex specific Glomerular Filtration Rate in the Abu Dhabi population and its association with mortality and Atherosclerotic cardiovascular outcome. A Retrospective Cohort Study": Table 1

Table 1 Subjects’ characteristics

| ADRS 2011-2013 cohort | | |
| --- | --- | --- |
|  | Mean | SD |
| BMI | 28.8 | 6.3 |
| GFR | 111.3 | 18.6 |
| SBP | 120.5 | 15.2 |
| DBP | 73.5 | 10.5 |
| Total cholesterol | 4.8 | 2.1 |
| HBA1C | 5.9 | 1.1 |
| HDL | 1.3 | 0.3 |
| Age | 38.9 | 15 |
| Prevelance of Chronic Diseases | | |
| Diabetes Mellitus | 22.2% | |
| Hypertension | 19.3% | |
| Smoking among males | 16.30% | |
