## Appendix 1 for "Estimation of age and sex specific Glomerular Filtration Rate in the Abu Dhabi population and its association with mortality and Atherosclerotic cardiovascular outcome. A Retrospective Cohort Study"

| Females | | | | | | | | | |
|---|---|---|---|---|---|---|---|---|---|
| Age | Gender | Percent3 | Percent10 | Percent25 | median | Percent75 | Percent90 | Percent95 | Percent97 |
| 14 | 1.00 | 110.98 | 122.62 | 130.65 | 137.13 | 142.04 | 145.58 | 147.40 | 148.49 |
| 15 | 1.00 | 110.63 | 121.82 | 129.72 | 136.19 | 141.15 | 144.75 | 146.61 | 147.72 |
| 16 | 1.00 | 110.24 | 121.02 | 128.80 | 135.26 | 140.26 | 143.91 | 145.81 | 146.94 |
| 17 | 1.00 | 109.81 | 120.21 | 127.88 | 134.33 | 139.38 | 143.08 | 145.01 | 146.17 |
| 18 | 1.00 | 109.34 | 119.40 | 126.96 | 133.40 | 138.49 | 142.25 | 144.22 | 145.40 |
| 19 | 1.00 | 108.86 | 118.59 | 126.04 | 132.48 | 137.61 | 141.42 | 143.42 | 144.63 |
| 20 | 1.00 | 108.36 | 117.80 | 125.14 | 131.57 | 136.73 | 140.60 | 142.64 | 143.86 |
| 21 | 1.00 | 107.83 | 117.00 | 124.25 | 130.67 | 135.87 | 139.78 | 141.85 | 143.10 |
| 22 | 1.00 | 107.28 | 116.21 | 123.36 | 129.77 | 135.00 | 138.96 | 141.06 | 142.34 |
| 23 | 1.00 | 106.68 | 115.39 | 122.46 | 128.85 | 134.11 | 138.12 | 140.26 | 141.55 |
| 24 | 1.00 | 106.02 | 114.54 | 121.54 | 127.92 | 133.21 | 137.27 | 139.43 | 140.75 |
| 25 | 1.00 | 105.33 | 113.68 | 120.60 | 126.97 | 132.29 | 136.39 | 138.59 | 139.93 |
| 26 | 1.00 | 104.61 | 112.79 | 119.66 | 126.02 | 131.37 | 135.50 | 137.73 | 139.09 |
| 27 | 1.00 | 103.87 | 111.91 | 118.72 | 125.07 | 130.44 | 134.61 | 136.87 | 138.24 |
| 28 | 1.00 | 103.14 | 111.04 | 117.79 | 124.12 | 129.52 | 133.73 | 136.01 | 137.40 |
| 29 | 1.00 | 102.40 | 110.18 | 116.87 | 123.19 | 128.60 | 132.84 | 135.14 | 136.55 |
| 30 | 1.00 | 101.65 | 109.33 | 115.96 | 122.27 | 127.69 | 131.96 | 134.28 | 135.70 |
| 31 | 1.00 | 100.88 | 108.47 | 115.06 | 121.36 | 126.79 | 131.07 | 133.41 | 134.84 |
| 32 | 1.00 | 100.10 | 107.62 | 114.17 | 120.46 | 125.89 | 130.19 | 132.53 | 133.97 |
| 33 | 1.00 | 99.31 | 106.77 | 113.29 | 119.56 | 124.99 | 129.29 | 131.65 | 133.09 |
| 34 | 1.00 | 98.49 | 105.91 | 112.41 | 118.66 | 124.09 | 128.39 | 130.75 | 132.19 |
| 35 | 1.00 | 97.65 | 105.04 | 111.52 | 117.76 | 123.18 | 127.48 | 129.84 | 131.28 |
| 36 | 1.00 | 96.80 | 104.18 | 110.64 | 116.86 | 122.27 | 126.56 | 128.91 | 130.36 |
| 37 | 1.00 | 95.94 | 103.31 | 109.76 | 115.97 | 121.36 | 125.64 | 127.98 | 129.42 |
| 38 | 1.00 | 95.07 | 102.45 | 108.90 | 115.09 | 120.46 | 124.72 | 127.05 | 128.48 |
| 39 | 1.00 | 94.19 | 101.60 | 108.04 | 114.22 | 119.56 | 123.80 | 126.11 | 127.53 |
| 40 | 1.00 | 93.31 | 100.75 | 107.20 | 113.35 | 118.67 | 122.88 | 125.17 | 126.58 |
| 41 | 1.00 | 92.44 | 99.91 | 106.36 | 112.50 | 117.79 | 121.96 | 124.23 | 125.63 |
| 42 | 1.00 | 91.59 | 99.09 | 105.55 | 111.67 | 116.92 | 121.05 | 123.30 | 124.68 |
| 43 | 1.00 | 90.74 | 98.28 | 104.74 | 110.83 | 116.05 | 120.14 | 122.37 | 123.74 |
| 44 | 1.00 | 89.89 | 97.48 | 103.93 | 110.00 | 115.18 | 119.23 | 121.43 | 122.78 |
| 45 | 1.00 | 89.05 | 96.67 | 103.12 | 109.16 | 114.30 | 118.32 | 120.50 | 121.83 |
| 46 | 1.00 | 88.22 | 95.87 | 102.32 | 108.33 | 113.43 | 117.40 | 119.56 | 120.87 |
| 47 | 1.00 | 87.41 | 95.09 | 101.52 | 107.50 | 112.56 | 116.49 | 118.62 | 119.92 |
| 48 | 1.00 | 86.61 | 94.31 | 100.73 | 106.67 | 111.69 | 115.58 | 117.69 | 118.97 |
| 49 | 1.00 | 85.82 | 93.53 | 99.93 | 105.84 | 110.81 | 114.66 | 116.75 | 118.02 |
| 50 | 1.00 | 85.03 | 92.74 | 99.12 | 105.00 | 109.92 | 113.74 | 115.80 | 117.06 |
| 51 | 1.00 | 84.25 | 91.96 | 98.31 | 104.15 | 109.03 | 112.81 | 114.85 | 116.09 |
| 52 | 1.00 | 83.48 | 91.18 | 97.50 | 103.29 | 108.13 | 111.87 | 113.89 | 115.12 |
| 53 | 1.00 | 82.71 | 90.39 | 96.68 | 102.42 | 107.22 | 110.93 | 112.92 | 114.14 |
| 54 | 1.00 | 81.94 | 89.59 | 95.85 | 101.55 | 106.30 | 109.97 | 111.95 | 113.15 |
| 55 | 1.00 | 81.18 | 88.80 | 95.01 | 100.67 | 105.38 | 109.01 | 110.96 | 112.16 |
| 56 | 1.00 | 80.44 | 88.02 | 94.18 | 99.79 | 104.45 | 108.05 | 109.98 | 111.16 |
| 57 | 1.00 | 79.72 | 87.25 | 93.36 | 98.92 | 103.54 | 107.10 | 109.01 | 110.18 |
| 58 | 1.00 | 79.03 | 86.50 | 92.56 | 98.06 | 102.63 | 106.16 | 108.05 | 109.21 |
| 59 | 1.00 | 78.38 | 85.78 | 91.77 | 97.22 | 101.74 | 105.23 | 107.10 | 108.25 |
| 60 | 1.00 | 77.74 | 85.06 | 91.00 | 96.38 | 100.86 | 104.31 | 106.16 | 107.30 |
| 61 | 1.00 | 77.13 | 84.37 | 90.23 | 95.56 | 99.98 | 103.39 | 105.23 | 106.35 |

| | | | | | | | | | |
|---|---|---|---|---|---|---|---|---|---|
| 62 | 1.00 | 76.54 | 83.69 | 89.48 | 94.74 | 99.11 | 102.49 | 104.30 | 105.41 |
| 63 | 1.00 | 75.97 | 83.01 | 88.72 | 93.92 | 98.24 | 101.58 | 103.38 | 104.47 |
| 64 | 1.00 | 75.40 | 82.34 | 87.97 | 93.10 | 97.37 | 100.67 | 102.45 | 103.53 |
| 65 | 1.00 | 74.83 | 81.66 | 87.21 | 92.27 | 96.49 | 99.75 | 101.51 | 102.58 |
| 66 | 1.00 | 74.27 | 80.98 | 86.45 | 91.44 | 95.60 | 98.82 | 100.56 | 101.62 |
| 67 | 1.00 | 73.69 | 80.29 | 85.68 | 90.59 | 94.70 | 97.89 | 99.60 | 100.65 |
| 68 | 1.00 | 73.11 | 79.59 | 84.89 | 89.73 | 93.79 | 96.93 | 98.63 | 99.67 |
| 69 | 1.00 | 72.51 | 78.87 | 84.08 | 88.85 | 92.85 | 95.95 | 97.63 | 98.66 |
| 70 | 1.00 | 71.89 | 78.13 | 83.24 | 87.94 | 91.89 | 94.95 | 96.61 | 97.62 |
| 71 | 1.00 | 71.26 | 77.36 | 82.39 | 87.01 | 90.90 | 93.93 | 95.56 | 96.57 |
| 72 | 1.00 | 70.60 | 76.58 | 81.51 | 86.06 | 89.89 | 92.87 | 94.49 | 95.48 |
| 73 | 1.00 | 69.91 | 75.76 | 80.60 | 85.07 | 88.85 | 91.79 | 93.39 | 94.37 |
| 74 | 1.00 | 69.21 | 74.93 | 79.67 | 84.06 | 87.78 | 90.68 | 92.26 | 93.23 |
| 75 | 1.00 | 68.49 | 74.08 | 78.72 | 83.03 | 86.69 | 89.55 | 91.11 | 92.07 |
| 76 | 1.00 | 67.76 | 73.20 | 77.75 | 81.98 | 85.58 | 88.40 | 89.94 | 90.88 |
| 77 | 1.00 | 67.01 | 72.32 | 76.76 | 80.91 | 84.45 | 87.22 | 88.74 | 89.67 |
| 78 | 1.00 | 66.25 | 71.41 | 75.75 | 79.82 | 83.29 | 86.03 | 87.52 | 88.44 |
| 79 | 1.00 | 65.47 | 70.49 | 74.73 | 78.71 | 82.12 | 84.81 | 86.28 | 87.18 |
| 80 | 1.00 | 64.68 | 69.55 | 73.69 | 77.58 | 80.93 | 83.57 | 85.02 | 85.91 |
| 81 | 1.00 | 63.88 | 68.60 | 72.63 | 76.44 | 79.71 | 82.31 | 83.73 | 84.61 |
| 82 | 1.00 | 63.07 | 67.65 | 71.56 | 75.28 | 78.49 | 81.04 | 82.44 | 83.30 |
| 83 | 1.00 | 62.25 | 66.68 | 70.48 | 74.11 | 77.25 | 79.75 | 81.12 | 81.97 |
| 84 | 1.00 | 61.42 | 65.70 | 69.40 | 72.93 | 76.00 | 78.44 | 79.79 | 80.62 |
| 85 | 1.00 | 60.59 | 64.72 | 68.30 | 71.74 | 74.73 | 77.13 | 78.45 | 79.27 |
| 86 | 1.00 | 59.76 | 63.74 | 67.20 | 70.54 | 73.46 | 75.80 | 77.10 | 77.90 |
| 87 | 1.00 | 58.93 | 62.75 | 66.10 | 69.34 | 72.18 | 74.47 | 75.73 | 76.52 |
| 88 | 1.00 | 58.10 | 61.77 | 64.99 | 68.13 | 70.89 | 73.12 | 74.36 | 75.13 |
| 89 | 1.00 | 57.27 | 60.78 | 63.89 | 66.92 | 69.61 | 71.78 | 72.98 | 73.73 |

| Males | | | | | | | | | |
|---|---|---|---|---|---|---|---|---|---|
| Age | Gender | Percent3 | Percent10 | Percent25 | median | Percent75 | Percent90 | Percent95 | Percent97 |
| 14 | 2.00 | 111.13 | 119.69 | 127.40 | 134.98 | 141.65 | 146.99 | 149.91 | 151.70 |
| 15 | 2.00 | 109.69 | 118.33 | 126.12 | 133.76 | 140.49 | 145.87 | 148.81 | 150.62 |
| 16 | 2.00 | 108.26 | 116.99 | 124.84 | 132.55 | 139.33 | 144.74 | 147.71 | 149.53 |
| 17 | 2.00 | 106.84 | 115.65 | 123.57 | 131.33 | 138.16 | 143.61 | 146.60 | 148.43 |
| 18 | 2.00 | 105.43 | 114.32 | 122.30 | 130.12 | 136.99 | 142.48 | 145.48 | 147.33 |
| 19 | 2.00 | 104.03 | 112.99 | 121.03 | 128.90 | 135.82 | 141.34 | 144.37 | 146.22 |
| 20 | 2.00 | 102.65 | 111.68 | 119.77 | 127.70 | 134.65 | 140.21 | 143.25 | 145.12 |
| 21 | 2.00 | 101.30 | 110.39 | 118.53 | 126.50 | 133.50 | 139.08 | 142.13 | 144.01 |
| 22 | 2.00 | 99.98 | 109.13 | 117.31 | 125.32 | 132.35 | 137.95 | 141.02 | 142.91 |
| 23 | 2.00 | 98.69 | 107.89 | 116.12 | 124.16 | 131.21 | 136.84 | 139.92 | 141.82 |
| 24 | 2.00 | 97.44 | 106.69 | 114.95 | 123.02 | 130.10 | 135.74 | 138.83 | 140.73 |
| 25 | 2.00 | 96.22 | 105.52 | 113.82 | 121.91 | 129.01 | 134.66 | 137.75 | 139.65 |
| 26 | 2.00 | 95.04 | 104.39 | 112.72 | 120.83 | 127.94 | 133.59 | 136.69 | 138.59 |
| 27 | 2.00 | 93.90 | 103.30 | 111.65 | 119.78 | 126.89 | 132.54 | 135.64 | 137.54 |
| 28 | 2.00 | 92.81 | 102.25 | 110.63 | 118.76 | 125.87 | 131.51 | 134.60 | 136.50 |
| 29 | 2.00 | 91.75 | 101.24 | 109.64 | 117.78 | 124.87 | 130.50 | 133.58 | 135.47 |
| 30 | 2.00 | 90.74 | 100.27 | 108.69 | 116.82 | 123.90 | 129.51 | 132.58 | 134.46 |
| 31 | 2.00 | 89.78 | 99.34 | 107.77 | 115.90 | 122.96 | 128.54 | 131.59 | 133.46 |
| 32 | 2.00 | 88.85 | 98.46 | 106.89 | 115.00 | 122.03 | 127.59 | 130.62 | 132.48 |
| 33 | 2.00 | 87.96 | 97.60 | 106.04 | 114.13 | 121.13 | 126.65 | 129.66 | 131.50 |
| 34 | 2.00 | 87.10 | 96.78 | 105.21 | 113.29 | 120.25 | 125.73 | 128.71 | 130.53 |
| 35 | 2.00 | 86.26 | 95.98 | 104.42 | 112.46 | 119.38 | 124.81 | 127.77 | 129.58 |
| 36 | 2.00 | 85.45 | 95.21 | 103.64 | 111.65 | 118.52 | 123.91 | 126.84 | 128.63 |
| 37 | 2.00 | 84.66 | 94.45 | 102.88 | 110.85 | 117.67 | 123.02 | 125.91 | 127.68 |
| 38 | 2.00 | 83.89 | 93.71 | 102.13 | 110.06 | 116.83 | 122.12 | 124.98 | 126.73 |
| 39 | 2.00 | 83.12 | 92.98 | 101.39 | 109.29 | 116.00 | 121.23 | 124.06 | 125.79 |
| 40 | 2.00 | 82.37 | 92.27 | 100.66 | 108.52 | 115.17 | 120.35 | 123.14 | 124.85 |
| 41 | 2.00 | 81.64 | 91.57 | 99.95 | 107.76 | 114.36 | 119.48 | 122.24 | 123.92 |
| 42 | 2.00 | 80.93 | 90.90 | 99.26 | 107.02 | 113.55 | 118.61 | 121.34 | 123.00 |
| 43 | 2.00 | 80.25 | 90.24 | 98.58 | 106.29 | 112.76 | 117.76 | 120.45 | 122.09 |
| 44 | 2.00 | 79.59 | 89.61 | 97.92 | 105.58 | 111.98 | 116.92 | 119.58 | 121.19 |
| 45 | 2.00 | 78.97 | 88.99 | 97.28 | 104.88 | 111.22 | 116.10 | 118.72 | 120.31 |
| 46 | 2.00 | 78.36 | 88.40 | 96.65 | 104.19 | 110.47 | 115.29 | 117.87 | 119.45 |
| 47 | 2.00 | 77.78 | 87.82 | 96.04 | 103.52 | 109.72 | 114.49 | 117.04 | 118.59 |
| 48 | 2.00 | 77.21 | 87.25 | 95.43 | 102.85 | 108.99 | 113.70 | 116.22 | 117.75 |
| 49 | 2.00 | 76.66 | 86.69 | 94.83 | 102.18 | 108.26 | 112.91 | 115.40 | 116.91 |
| 50 | 2.00 | 76.12 | 86.14 | 94.23 | 101.52 | 107.54 | 112.14 | 114.59 | 116.09 |
| 51 | 2.00 | 75.58 | 85.59 | 93.63 | 100.87 | 106.82 | 111.37 | 113.79 | 115.27 |
| 52 | 2.00 | 75.05 | 85.03 | 93.03 | 100.21 | 106.10 | 110.60 | 113.00 | 114.45 |
| 53 | 2.00 | 74.51 | 84.48 | 92.43 | 99.55 | 105.39 | 109.83 | 112.20 | 113.64 |
| 54 | 2.00 | 73.99 | 83.93 | 91.84 | 98.90 | 104.68 | 109.08 | 111.42 | 112.84 |
| 55 | 2.00 | 73.46 | 83.38 | 91.25 | 98.25 | 103.98 | 108.33 | 110.65 | 112.05 |
| 56 | 2.00 | 72.94 | 82.84 | 90.66 | 97.61 | 103.28 | 107.58 | 109.88 | 111.27 |
| 57 | 2.00 | 72.42 | 82.29 | 90.07 | 96.96 | 102.58 | 106.85 | 109.12 | 110.49 |
| 58 | 2.00 | 71.90 | 81.74 | 89.47 | 96.32 | 101.89 | 106.11 | 108.36 | 109.72 |
| 59 | 2.00 | 71.36 | 81.18 | 88.87 | 95.66 | 101.19 | 105.37 | 107.60 | 108.95 |
| 60 | 2.00 | 70.82 | 80.62 | 88.26 | 95.00 | 100.48 | 104.63 | 106.83 | 108.17 |
| 61 | 2.00 | 70.26 | 80.03 | 87.64 | 94.33 | 99.77 | 103.87 | 106.06 | 107.38 |

| | | | | | | | | | |
|---|---|---|---|---|---|---|---|---|---|
| 62 | 2.00 | 69.70 | 79.44 | 87.01 | 93.65 | 99.04 | 103.11 | 105.28 | 106.59 |
| 63 | 2.00 | 69.12 | 78.83 | 86.36 | 92.96 | 98.31 | 102.34 | 104.49 | 105.79 |
| 64 | 2.00 | 68.53 | 78.22 | 85.70 | 92.26 | 97.56 | 101.56 | 103.69 | 104.98 |
| 65 | 2.00 | 67.93 | 77.59 | 85.03 | 91.54 | 96.80 | 100.77 | 102.88 | 104.16 |
| 66 | 2.00 | 67.32 | 76.94 | 84.34 | 90.81 | 96.03 | 99.97 | 102.06 | 103.33 |
| 67 | 2.00 | 66.70 | 76.29 | 83.65 | 90.07 | 95.25 | 99.15 | 101.23 | 102.48 |
| 68 | 2.00 | 66.08 | 75.63 | 82.95 | 89.32 | 94.46 | 98.33 | 100.39 | 101.63 |
| 69 | 2.00 | 65.46 | 74.97 | 82.23 | 88.56 | 93.66 | 97.50 | 99.54 | 100.78 |
| 70 | 2.00 | 64.84 | 74.30 | 81.52 | 87.80 | 92.85 | 96.66 | 98.69 | 99.91 |
| 71 | 2.00 | 64.21 | 73.62 | 80.79 | 87.02 | 92.04 | 95.82 | 97.83 | 99.04 |
| 72 | 2.00 | 63.58 | 72.94 | 80.06 | 86.24 | 91.22 | 94.97 | 96.96 | 98.16 |
| 73 | 2.00 | 62.94 | 72.24 | 79.31 | 85.45 | 90.39 | 94.10 | 96.08 | 97.27 |
| 74 | 2.00 | 62.30 | 71.54 | 78.56 | 84.65 | 89.55 | 93.23 | 95.19 | 96.38 |
| 75 | 2.00 | 61.66 | 70.84 | 77.80 | 83.84 | 88.70 | 92.36 | 94.30 | 95.47 |
| 76 | 2.00 | 61.01 | 70.13 | 77.04 | 83.03 | 87.84 | 91.47 | 93.40 | 94.56 |
| 77 | 2.00 | 60.37 | 69.42 | 76.27 | 82.21 | 86.98 | 90.58 | 92.49 | 93.65 |
| 78 | 2.00 | 59.74 | 68.71 | 75.50 | 81.39 | 86.12 | 89.69 | 91.58 | 92.73 |
| 79 | 2.00 | 59.11 | 68.00 | 74.73 | 80.56 | 85.25 | 88.79 | 90.67 | 91.81 |
| 80 | 2.00 | 58.49 | 67.29 | 73.96 | 79.73 | 84.38 | 87.89 | 89.75 | 90.88 |
