## Supplementary figures and images for "Estimation of age and sex specific Glomerular Filtration Rate in the Abu Dhabi population and its association with mortality and Atherosclerotic cardiovascular outcome. A Retrospective Cohort Study"

### Appendix 2

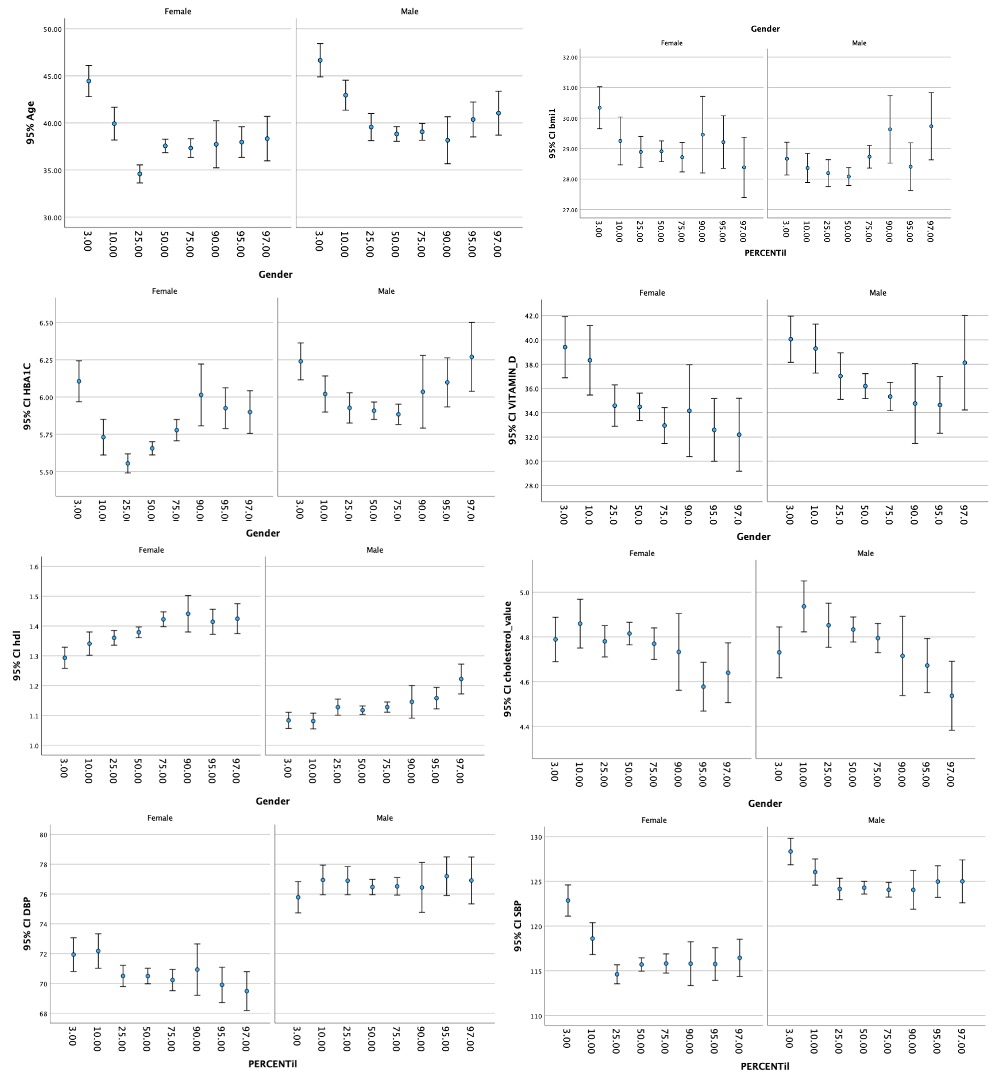
